# Relationship Between Physiological Mirror Activity and Corticomuscular Coherence During a Finger Dexterity Task Among Healthy Young and Older Adults

**DOI:** 10.64898/2026.08.12.26360287

**Authors:** Shun Sawai, Shin Murata, Naoki Shimizu, Shoya Fujikawa, Ryosuke Yamamoto, Takato Nishida, Yusuke Shizuka, Hideki Nakano

## Abstract

Physiological mirror activity (pMA) is the increase in involuntary muscle activity observed on the contralateral side during unilateral voluntary movement in neurologically healthy participants. This cross-sectional study aimed to explore the relationship between pMA and corticomuscular coherence (CMC) during finger dexterity tasks in young and older adults. Thirty-one right-handed young adults and 24 older adults performed a left-hand finger dexterity task. Electroencephalogram (EEG) signals were recorded from C3 and C4, and electromyogram (EMG) signals were collected from bilateral finger flexors and extensors. pMA was quantified as the change in right-hand EMG from rest to task. Gamma-band CMC was calculated from task-related EEG–EMG pairs, and its association with pMA was analyzed. In young adults, greater pMA was associated with lower CMC (C3– and C4-right flexors), whereas in older adults, greater pMA was associated with higher CMC (C3-left flexor). Young adults may suppress pMA emergence by appropriately monitoring and inhibiting activity, in the hand not performing the task. Conversely, in older adults, the mobilization of the ipsilateral motor cortex may have contributed to pMA emergence. This study suggests that the neuromuscular mechanisms involved in pMA during finger dexterity tasks differ between young and older adults.

## Introduction

Mirror activity (MA) is an increase in involuntary muscle activity on the contralateral side that occurs during voluntary movement on the ipsilateral side [1]. MA appears prominently in neurological disorders, including stroke [2] and Parkinson’s disease [3], and this is termed pathological MA. Such prominent MA impairs independent bilateral movement during activities of daily living, prompting research into methods for evaluating MA and interventions to suppress it. On the other hand, mild MA is observed in neurologically healthy individuals, termed physiological MA (pMA). pMA is strongly evident in children, diminishes during adolescence with development, but reappears strongly in older adults with aging [4]. Therefore, strong pMA in older adults may be associated with a decline in motor function and linked to healthy aging [5]. Furthermore, regarding pMA neural mechanisms, the active and passive hands may share the same neural origin [6]. This is attributed to the activation of both cerebral hemispheres due to a reduction in interhemispheric inhibitory function mediated by the corpus callosum [7]. Reportedly, the dorsal premotor areas may be involved in pMA; repetitive transcranial magnetic stimulation [8,9] and transcranial direct current stimulation [10] applied to the dorsal premotor areas can alter pMA. This has led to an increase in the investigations of neural mechanisms of pMA. However, the neuromuscular mechanisms underlying pMA remain unclear.

Corticomuscular coherence (CMC), a correlation between the electroencephalogram (EEG) and electromyogram (EMG), serves as a neurophysiological indicator reflecting the functional connectivity between the central nervous system and peripheral muscles [11,12]. CMC reflects not only descending neural transmission from the brain to muscles but also ascending neural transmission from muscles to the brain [13]. Consequently, CMC is used as a parameter related to motor control and motor learning. Regarding the frequency response of CMCs, beta band CMCs are involved in low-intensity steady-state motor output, as they contribute to output stabilization and reduction of motor noise during voluntary movement, through both ascending and descending neural transmission [14,15]. Conversely, gamma-band CMCs are involved in dynamic motor control [16] and high-intensity steady-state motor output [17]. In addition, gamma-band CMCs are low during dynamic motor control in patients with afferent pathways blocked, and this may be influenced by proprioceptive input [18]. Thus, CMC can verify neuromuscular functions and mechanisms related to motor control, and is considered useful for understanding the neuromuscular mechanisms of pMA. Specifically, using gamma-band CMC may enable verification of pMA neuromuscular mechanisms during finger dexterity movements that require dynamic control.

Therefore, this study aimed to explore the relationship between pMA and CMC during finger dexterity tasks in healthy young and older adults. Our findings may enable us to propose new hypotheses regarding the neuromuscular mechanisms of pMA and its age-related changes.

## Results

First, the chi-square test revealed a significant sex ratio imbalance between the young and older adults (χ² = 6.58, *p* = 0.01). After adjusting for sex and comparing minimum RMS in the resting phase, no significant muscle × group interaction was observed (*F* = 1.06, *p* = 0.31, partial η^2^ = 0.02). A significant main effect of group was found (*F* = 4.87, *p* = 0.03, partial η^2^ = 0.09). Post-hoc tests showed that RMS was significantly higher in the older adults than in the young adults (*p* = 0.03). However, there was no significant main effect of muscle (*F* = 0.81, *p* = 0.37, partial η^2^ = 0.02) (Figure 4a). After adjusting for sex and comparing average RMS in the task phase, no significant muscle × group interaction (*F* = 1.60, *p* = 0.21, partial η^2^ = 0.03) was observed. A significant effect of group was found (*F* = 17.83, *p* < 0.01, partial η^2^ = 0.26). Post-hoc tests revealed that RMS was higher in the older adults than in the young adults (*p* < 0.01). However, there was no significant main effect of muscle (*F* = 0.89, *p* = 0.35, partial η^2^ = 0.02) (Figure 4b).

**Fig. 1.**
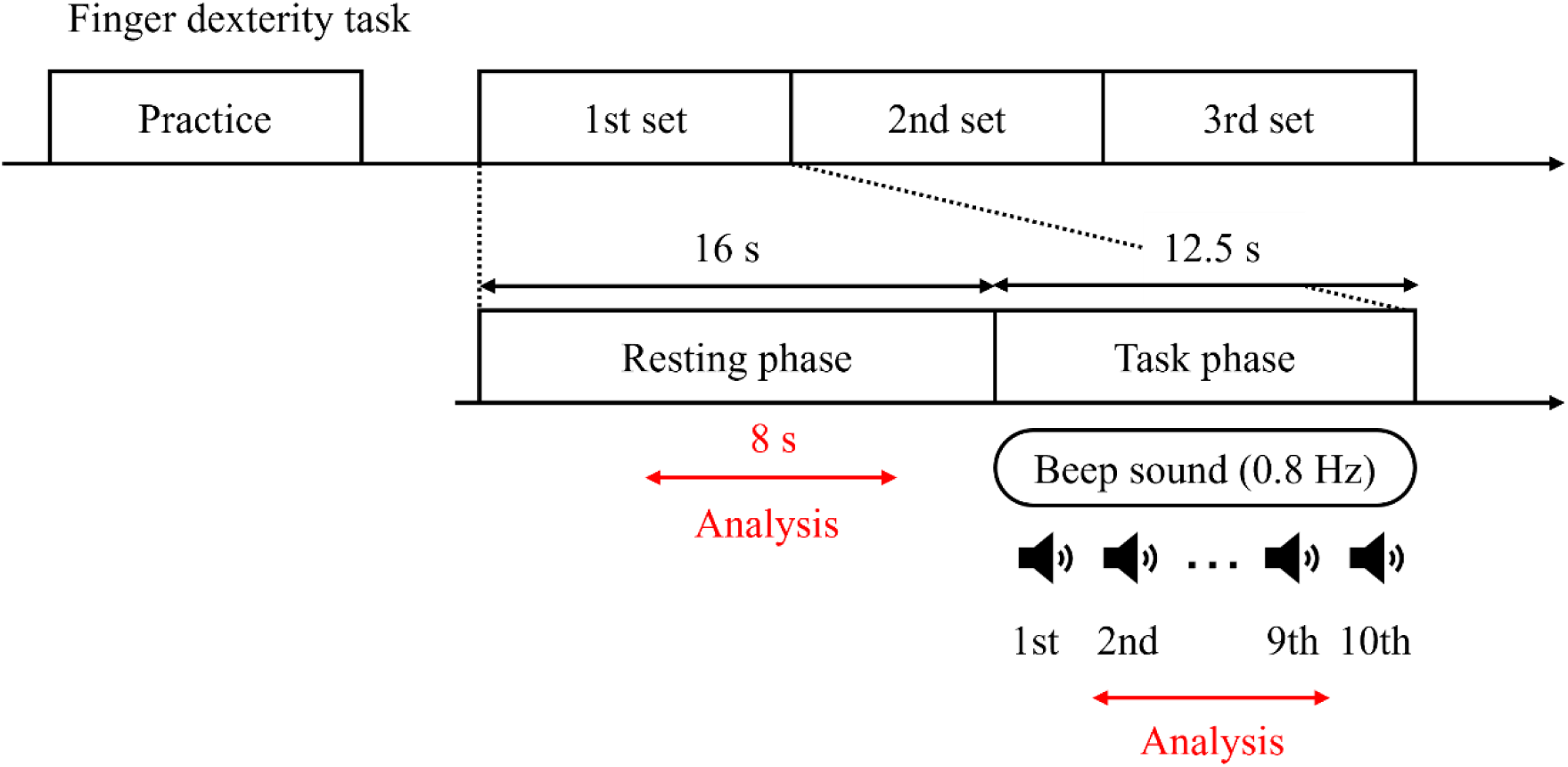
Study protocol. The finger dexterity task in this study comprised three sets. Each set had a 16-s resting phase and a 12.5-s task phase. During the resting phase, participants relaxed both upper limbs, while in the task phase, a 0.8 Hz beep sound was provided 10 times, and participants performed the finger dexterity task in sync with the beep sound. s, second.

**Fig. 2.**
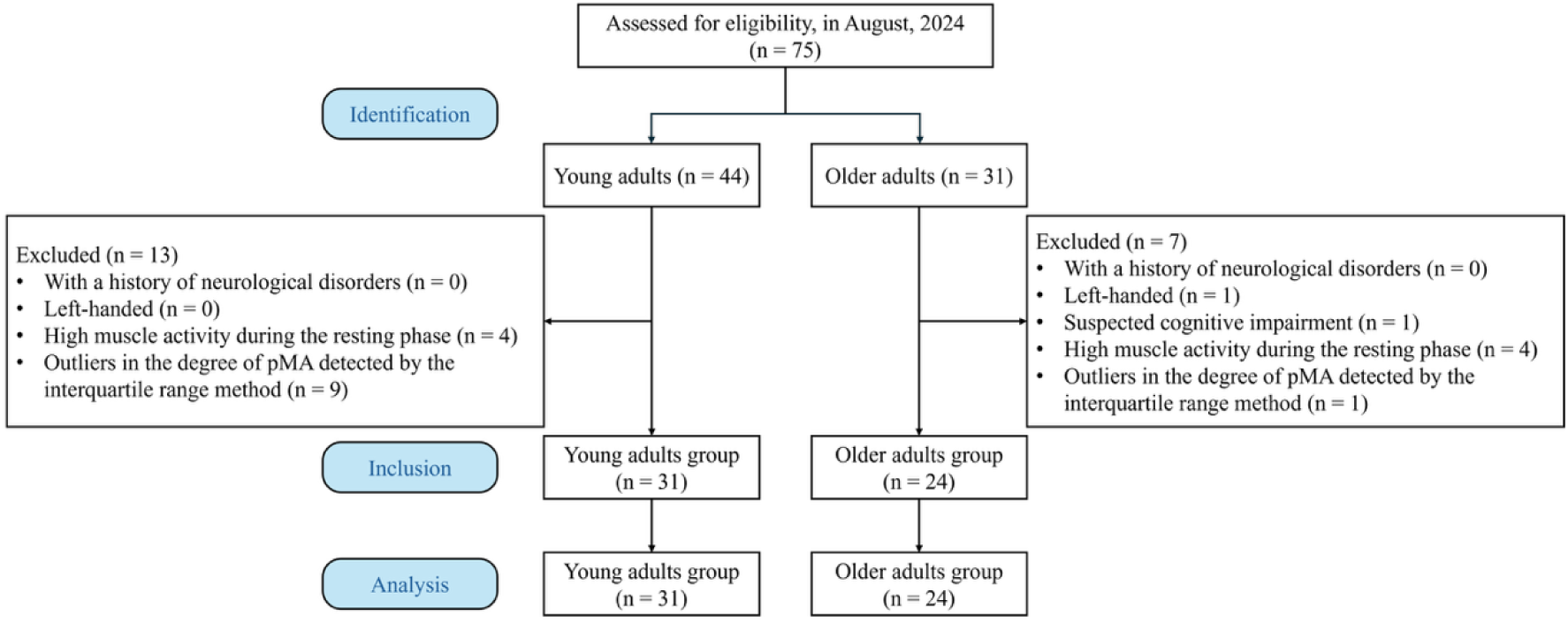
Participants’ inclusion flow chart. Forty-four young adults and 31 older adults were enrolled in the study. Thirty-one young adults and 24 older adults who met the inclusion criteria were included in the study and analyzed. pMA, physiological mirror activity.

**Fig. 3.**
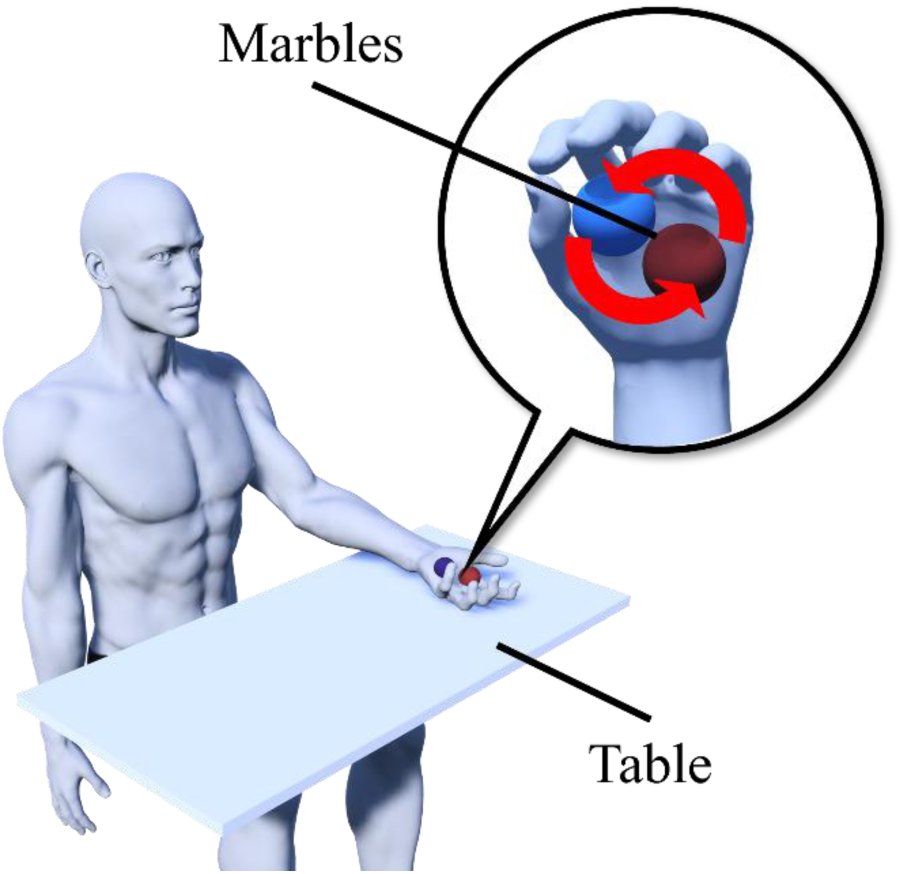
Finger dexterity task. Participants sat on chairs with backrests, relaxed their right arms at their sides, and placed their left forearms on the table. With their eyes closed, they rotated two marbles within their palms coordinated with a 0.8 Hz beep sound. The rotation speed was set to half a revolution per beep sound.

**Fig. 4.**
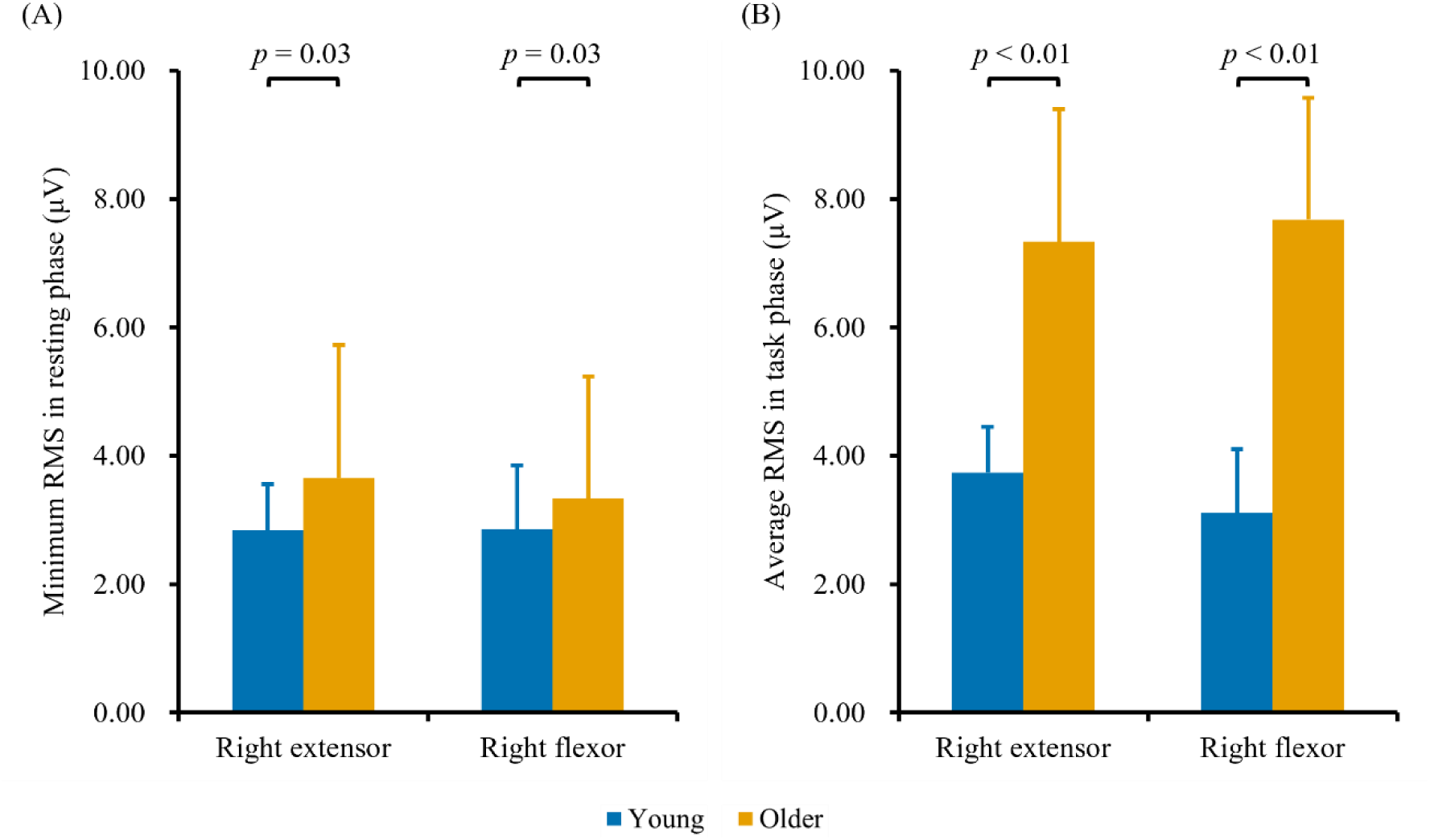
The comparison of the RMS in the right extensor and flexor. Young adults are represented by blue bars, and older adults by yellow bars. Error bars indicate standard deviation. (a) Comparison of the minimum RMS in the resting phase. The vertical axis shows the minimum RMS in the resting phase. RMS was significantly higher in older adults than in young adults (*p* = 0.03). (b) Comparison of average RMS in the task phase. The vertical axis shows average RMS in the task phase. RMS was significantly higher in the older adults than in the young adults (*p* < 0.01). RMS, root mean square; μV, microvolt.

Furthermore, when comparing the degree of pMA after adjusting for sex, no significant interaction was observed between the two factors of muscle × group (*F* = 1.37, *p* = 0.25, partial η^2^ = 0.03). In addition, the group factor showed a significant main effect (*F* = 28.31, *p* < 0.01, partial η^2^ = 0.35). Post-hoc tests showed that the degree of pMA was significantly higher in the older adults than in the young adults (*p* < 0.01). However, no significant main effect of the muscle factor was found (*F* = 0.69, *p* = 0.41, partial η^2^ = 0.01) (Figure 5). After adjusting for sex, the relationship between the degree of pMA and CMC was examined. In the young and older adults, the degree of pMA (right extensor) showed no significant correlation with any CMC (*p* > 0.05) (Figure 6). However, in the older adults, the degree of pMA (right flexor) showed a significant positive correlation with CMC (C3-left flexor) (partial ρ = 0.42, *p* = 0.049). Furthermore, in young adults, the degree of pMA (right flexor) showed a significant negative correlation with CMC (C3-right flexor) (partial ρ = −0.53, *p* < 0.01) and CMC (C4-right flexor) (partial ρ = −0.40, *p* = 0.03) (Figure 7).

**Fig. 5.**
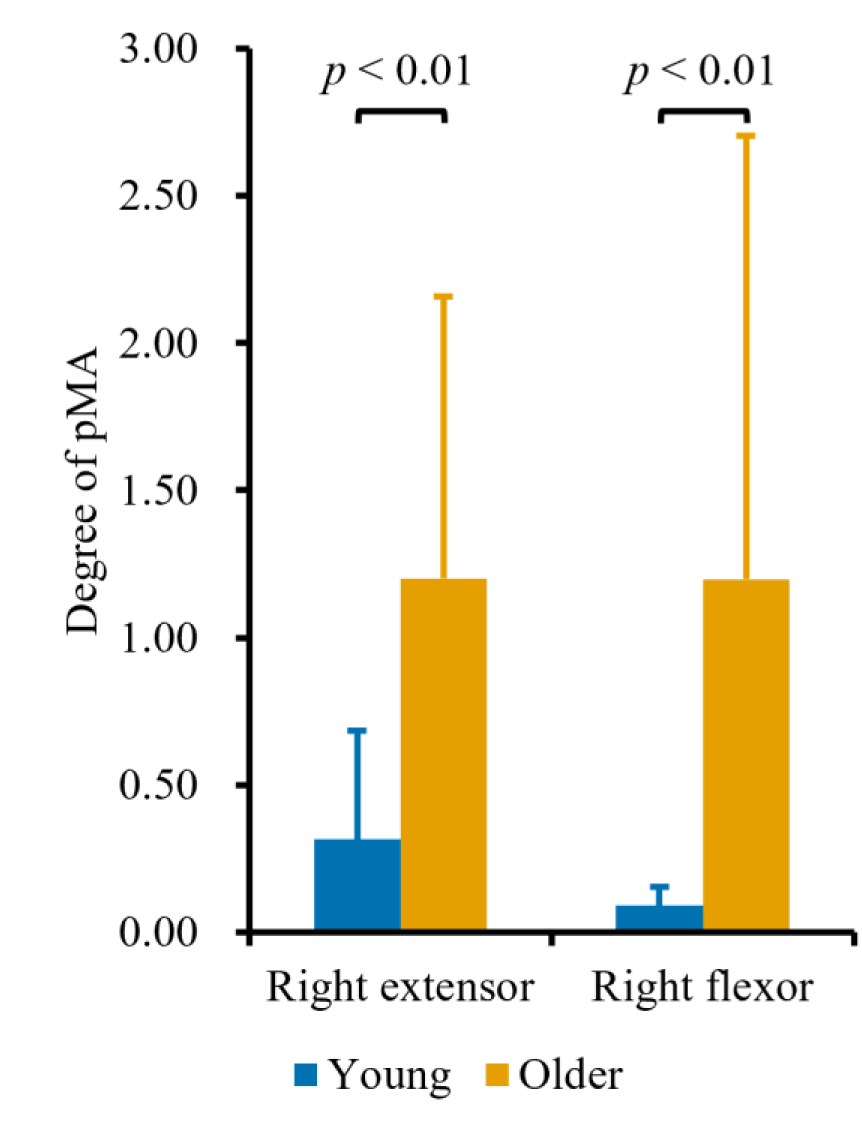
The comparison of the degree of pMA between young and older adults. The vertical axis represents the degree of pMA, and the error bars indicate the standard deviation. Young adults are represented by blue bars, and older adults by yellow bars. The degree of pMA was significantly higher in older adults than in young adults (*p* < 0.01). pMA, physiological mirror activity.

**Fig. 6.**
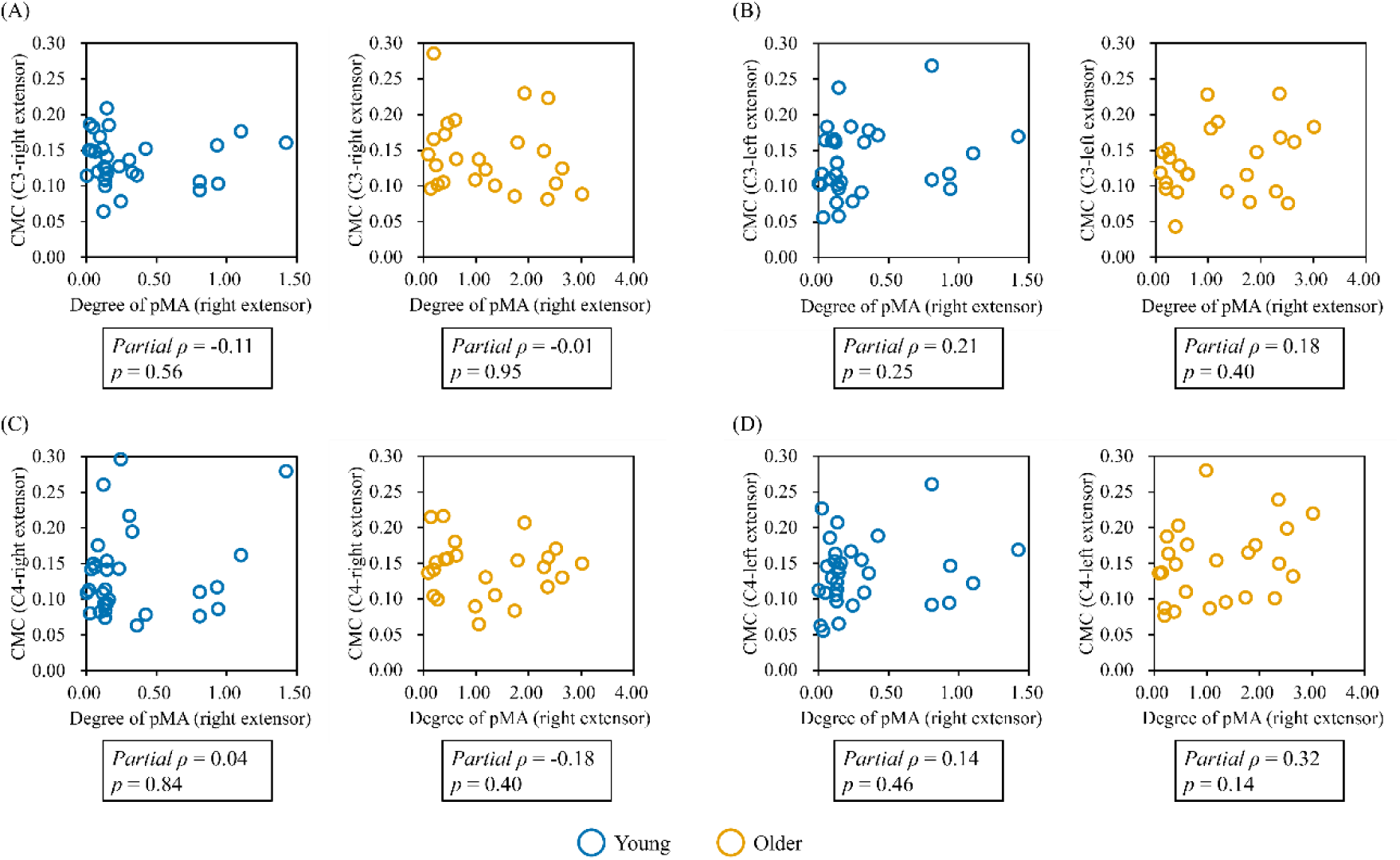
The relationship between the degree of pMA and CMC in the extensor. The scatter plot shows the relationship between the degree of pMA (right extensor) and CMC. Young adults are represented by blue plots, and older adults by yellow plots. (a) The relationship between the degree of pMA (right extensor) and CMC (C3-right extensor). No significant correlation was found in either young adults or older adults (*p* > 0.05). (b) The relationship between the degree of pMA (right extensor) and CMC (C3-left extensor). No significant correlation was found in either young or older adults (*p* > 0.05). (c) The relationship between the degree of pMA (right extensor) and CMC (C4-right extensor). No significant correlation was found in either young or older adults (*p* > 0.05). (d) The relationship between the degree of pMA (right extensor) and CMC (C4-left extensor). No significant correlation was found in either young or older adults (*p* > 0.05). CMC, corticomuscular coherence; pMA, physiological mirror activity.

**Fig. 7.**
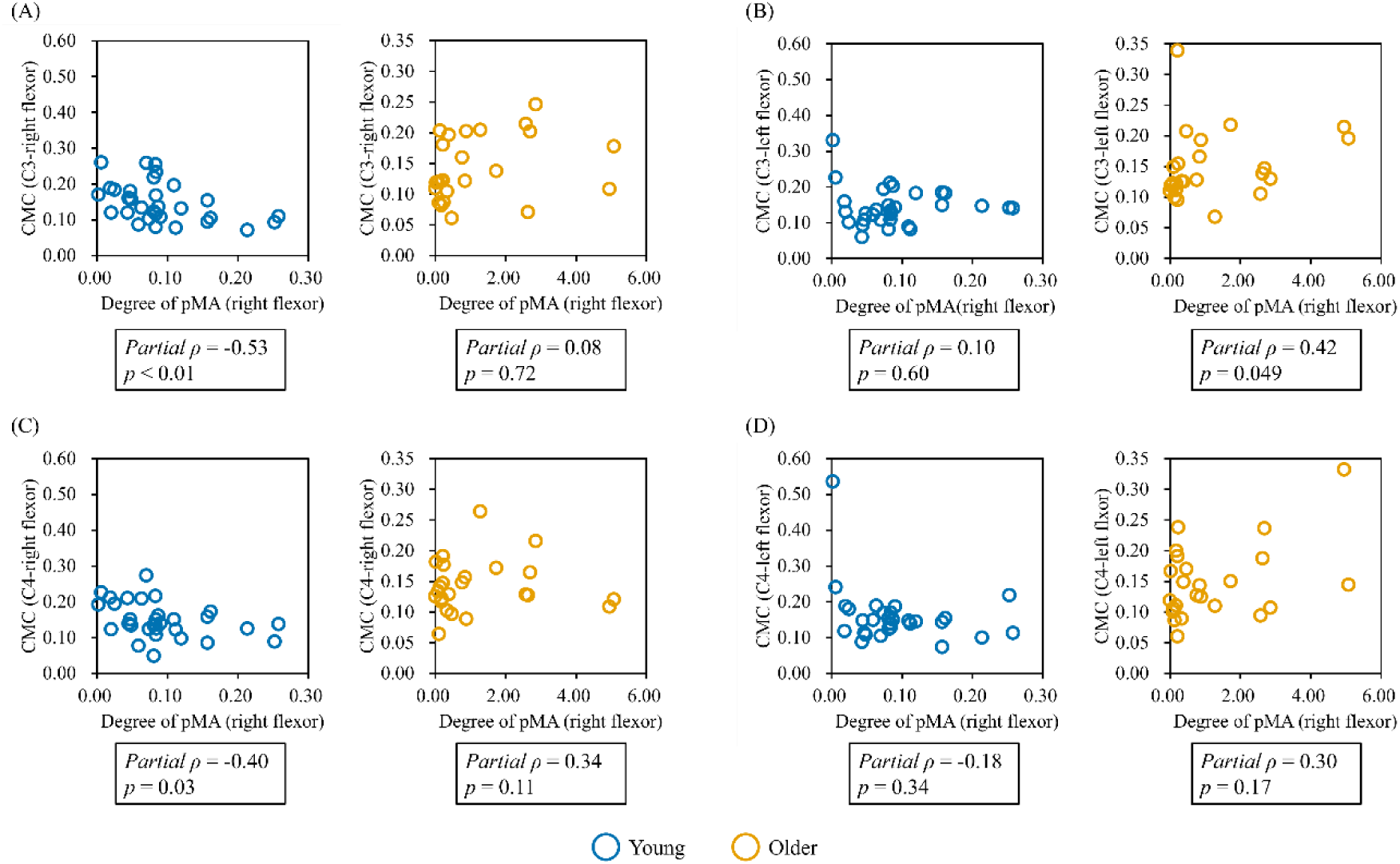
The relationship between the degree of pMA and CMC in the flexor. The scatter plot shows the relationship between the degree of pMA (right flexor) and CMC. Young adults are represented by blue plots, and older adults by yellow plots. (a) The relationship between the degree of pMA (right flexor) and CMC (C3-right flexor). In young adults, a significant negative correlation was observed between pMA (right flexor) and CMC (C3-right flexor) (*p* < 0.01). (b) The relationship between the degree of pMA (right flexor) and CMC (C3-left flexor). In older adults, a significant positive correlation was found between pMA (right flexor) and CMC (C3-left flexor) (*p* = 0.049). (c) The relationship between the degree of pMA (right flexor) and CMC (C4-right flexor). In young adults, a significant negative correlation was found between pMA (right flexor) and CMC (C4-right flexor) (*p* = 0.03). (d) The relationship between the degree of pMA (right flexor) and CMC (C4-left flexor). No significant correlation was found in either young or older adults (*p* > 0.05). CMC, corticomuscular coherence; pMA, physiological mirror activity.

## Discussion

This study explored the relationship between pMA and CMC during finger dexterity movements in healthy young adults and older adults. The results revealed that the degree of pMA was significantly higher in older adults than in young adults. Furthermore, in young adults, the degree of pMA (right flexor) showed a significant negative correlation with CMC (C3 and C4-right flexors). By contrast, the degree of pMA (right flexor) showed a significant positive correlation with CMC (C3-left flexor) among the older adults. These results suggest that the neurophysiological characteristics of the degree of pMA during finger dexterity movements may differ between young and older adults.

First, comparing the minimum RMS in the resting phase revealed that RMS was significantly higher in the older adults than in the young adults. This may be due to an age-related increase in background EMG. Reportedly, intracortical inhibitory function declines with aging, making it challenging for older adults to relax their muscles [19]. Furthermore, physiological tremors increase during rest with aging [20]. These reports indicate that older adults have difficulty maintaining muscle rest, suggesting the presence of factors that cause an increase in background EMG. Therefore, the minimum RMS in the resting phase may have been higher in older adults than in young adults in this study. The average RMS in the task phase was significantly higher in the older adults than in the young adults. This represented an increase in EMG activity due to performing finger dexterity exercises on the opposite side, which is pMA. This study suggests that the average RMS in the task phase for the right extensor and right flexor may have been higher in the older adults than in young adults, since pMA increases with age [4]. The degree of pMA was significantly higher in the older adults than in the young adults. This indicates that pMA is strongly present in older adults. pMA decreases rapidly with development from childhood to school age and increases again with aging from middle age onward. Thus, pMA exhibits a U-shaped appearance curve with age [4]. In this study, pMA appeared stronger in the older adults in their 70s than in the young adults in their 20s, consistent with previous research findings. Furthermore, pMA occurs strongly during high-effort exercise [21]. In addition, finger motor function declines with aging [22,23]. Therefore, the decline in finger motor function due to aging may have resulted in a relatively higher amount of effort required for the finger dexterity task among the older adults compared with the young adults in our study. This suggests that the degree of pMA was higher in the older adults than in the young adults.

An exploratory analysis of the relationship between the degree of pMA and CMC revealed a significant negative correlation between the degree of pMA (right flexor), CMC (C3-right flexor), and CMC (C4-right flexor) in young adults. This suggests that higher functional connectivity between C3-right flexor and C4-right flexor may be associated with smaller pMA in the right flexor. Gamma-band CMC is involved in dynamic motor control, reflecting the process of integrating multiple sensory inputs to generate subsequent motor programs [16]. Furthermore, gamma-band CMC is higher during isotonic exercise than during isometric exercise, indicating that proprioceptive input contributes to gamma-band CMC during dynamic motor control [24]. In this study, the higher CMC between the right flexor (not performing the finger dexterity task) and both motor cortices were associated with smaller pMA. This suggests that young adults with higher functional connectivity between the motor cortices of both hands and the hand not performing the task, who were effectively monitoring and suppressing activity in the non-task hand, may have exhibited smaller pMA.

Conversely, in the older adults, the degree of pMA (right flexor) and CMC (C3-left flexor) showed a significant positive correlation. This suggests that higher functional connectivity between C3-left flexor may have facilitated pMA in the right flexor. γ-band CMC reflects sensory integration and dynamic motor control [16,24]. Therefore, older adults may have attempted to compensate for sensory-motor integration function by enhancing γ-band functional connectivity between task-side muscles and the ipsilateral motor cortex during finger dexterity tasks. Furthermore, older adults exhibit reduced asymmetry between the left and right cerebral hemispheres, leading to bilateral activation [25,26]. In addition, studies report strong pMA in older adults with reduced interhemispheric inhibitory function between the left and right motor cortices [27]. These prior findings suggest that older adults may mobilize the ipsilateral motor cortex during unilateral movement, thereby generating pMA. In this study, stronger pMA was observed when functional connectivity between the muscles used during the task and the ipsilateral motor cortex was higher. Therefore, these results suggest that the mobilization of the ipsilateral motor cortex during finger dexterity tasks may be involved in the appearance of pMA in older adults.

Furthermore, a significant correlation between the degree of pMA and CMC was observed in flexors but not in extensors in our study. Reportedly, during palm-based object manipulation, finger flexors contribute to object movement, whereas finger extensors contribute to object stabilization [28]. This indicates that in palm-based object manipulation, finger flexors act as primary agonists and finger extensors as antagonist muscles. Therefore, the association between the degree of pMA and CMC may have been observed in flexors, which are directly involved in moving the grasped marble. Finger flexors exhibit higher cortical-muscle activity plasticity under conditions involving sensory input [29] than finger extensors. In this report, motor-evoked potentials before and after repetitive transcranial magnetic stimulation to the primary motor cortex showed no difference between finger extensors and flexors. However, changes in motor-evoked potentials before and after paired associative stimulation, involving peripheral sensory input, were substantially larger in finger flexors than in extensors. These results suggest that finger flexors exhibit high cortical-muscle activity plasticity mediated by sensory input. In this study, performing finger dexterity tasks requiring sensorimotor integration may have revealed the pMA-CMC relationship in flexors.

This study has some limitations. First, it is an exploratory study that comprehensively examines CMC related to pMA during finger dexterity movements across the left and right motor cortices, and the left and right extensor and flexor muscles. Future studies should conduct hypothesis-oriented research to localize the region of interest, enabling detailed examination of the proposed neuromuscular mechanism of pMA and its age-related changes. Second, this study is cross-sectional and cannot establish a causal relationship between pMA and CMC; therefore, future studies are warranted to clarify the causal relationship between pMA and CMC using non-invasive brain stimulation techniques such as transcranial magnetic stimulation or transcranial electrical stimulation.

This study explored the relationship between pMA and CMC during finger dexterity tasks in healthy young and older adults. Results showed that pMA was more pronounced in older adults than in young adults. Furthermore, in young adults, the degree of pMA exhibited a significant negative correlation with CMC (C3-right flexor) and CMC (C4-right flexor). Conversely, in the older adults, the degree of pMA (right flexor) showed a significant positive correlation with CMC (C3-left flexor). These results suggest that young adults may appropriately monitor and suppress activity in their hand not performing the task, potentially leading to a small pMA. By contrast, in older adults, the mobilization of the ipsilateral motor cortex may have contributed to the occurrence of pMA. This study suggests that the neuromuscular mechanisms involved in pMA during finger dexterity tasks differ between young and older adults.

## Methods

### Study protocol

In this study, participants performed three consecutive sets of finger dexterity tasks with their left hand after sufficient practice. Each set comprised a 16-s resting phase and a 12.5-s task phase. During the resting phase, participants were required to relax both upper limbs. In addition, an audio countdown of 3 s was provided at the end of the resting phase. During the task phase, a 0.8 Hz beep sound was emitted 10 times, and participants performed the finger dexterity task in sync with the beep sound (Figure 1). This study was conducted and reported in accordance with the Strengthening the Reporting of Observational Studies in Epidemiology (STROBE) statement for cross-sectional studies [30].

### Participants

This study included 44 healthy young adults (21 males, 23 females, age: 20.57 ± 1.58 years, height: 164.03 ± 7.32 cm, body weight: 57.18 ± 9.43 kg) and 31 community-dwelling older adults (5 males, 26 females, age: 74.84 ± 4.56 years, height: 155.82 ± 8.32 cm, body weight: 54.76 ± 10.14 kg) recruited in August 2024. Exclusion criteria for participants were: (i) participants with a history of neurological disorders such as cerebrovascular disease or neurodegenerative disease; (ii) participants confirmed to be left-handed by the Edinburgh Handedness Inventory [31]; (iii) participants considered cognitively impaired based on a score below 4 points on the Japanese version of the Rapid Dementia Screening Test [32]; (iv) participants with a degree of pMA below 0, indicating reduced muscle activity during task performance compared with rest performance; and (v) participants whose degree of pMA was judged an outlier, using the interquartile range (IQR) method (below the first quartile – 1.5 × IQR or above the third quartile + 1.5 × IQR). Conclusively, 31 young adults (12 males, 19 females, age: 20.45 ± 1.55 years, height: 163.31 ± 7.49 cm, body weight: 55.68 ± 7.52 kg) and 24 older adults (2 males, 22 females, age: 74.71 ± 5.09 years, height: 155.95 ± 8.24 cm, body weight: 53.92 ± 8.76 kg) were included in the analysis (Figure 2). This study was conducted in accordance with the Declaration of Helsinki and approved by the Ethics Committee of Kyoto Tachibana University (approval number: 24-83). Informed consent was obtained from all participants.

### Sample size calculation

Power analysis was performed using G*Power (G*Power 3.1; Heinrich Heine University, Düsseldorf, Germany) [33]. The power criteria in G*Power were set as follows: test family, F tests; analysis of covariance (ANCOVA): Fixed effects, main effects and interaction; effect size = 0.40 (Large); power (1–β error prob) = 0.80; α error prob = 0.05; numerator df = 1.00; number of groups = 2.00; number of covariates = 1.00. The results indicated that 52 participants were required.

### Finger dexterity task

All participants performed a finger dexterity task [34] involving rotating two marbles (diameter: 30 mm) counterclockwise with their left hands. Participants sat on chairs with backrests, relaxed their right arms at their sides, placed their left forearms on the table, and closed their eyes. Initially, all participants were given ample time to practice the task. Subsequently, a beep sound at a frequency of 0.8 Hz was generated, and participants were instructed to rotate the marbles half a turn per beep. Simultaneously, they were instructed to relax the right upper limb, not performing the task (Figure 3).

### Measurement

EEG and EMG data were recorded during the resting and task phases. EEG signals were obtained from the C3 and C4 electrodes, corresponding to the left motor-related area and right motor-related area, respectively, in the International 10–20 system, using active electrodes (AP-C151; Miyuki Giken Co., Ltd., Tokyo, Japan). EMG activity was recorded from the extensor digitorum muscles (extensor) and flexor digitorum superficialis muscles (flexor) on both sides using bipolar surface EMG electrodes (AP-C140; Miyuki Giken Co., Ltd., Tokyo, Japan). The reference electrode was placed on the left earlobe. Conductive gel was applied to both EEG and EMG electrodes to minimize electrical resistance. The EEG and EMG data were recorded at 2,000 Hz using a biological signal recording device (MP-6100; Miyuki Giken Co., Ltd., Tokyo, Japan). Furthermore, the resting phase was visually confirmed to lack any obvious muscle activity [35]. A ground electrode was used during data acquisition to reduce power supply noise.

### Data Analysis

The degree of pMA was calculated from the resting and task phase EMG data, and CMC was calculated from task phase EEG and EMG data. As part of data preprocessing, the EEG data underwent notch filtering (59–61 Hz), bandpass filtering (10–100 Hz), and down-sampling (1,000 Hz). The EMG data similarly underwent notch filtering (59–61 Hz), a bandpass filtering (10–500 Hz), and down-sampling (1,000 Hz). Rectification was not performed on either the EEG or EMG data in this study, owing to reports that rectification is not recommended for CMC analysis [36].

The degree of pMA was calculated by determining the root mean square (RMS) of the right extensor and right flexor during the resting phase and task phase. First, the analysis interval for the resting phase was set to 8 s, excluding the initial 5 s and final 3 s of this phase. The initial 5 s of the resting phase were excluded from the analysis interval to eliminate the influence of EMG activity from the preceding task phase. In addition, the final 3 s of the resting phase were excluded from the analysis interval because an audio countdown was provided before task initiation. Consequently, the total analysis time for the resting phase for each participant was 8 s × 3 sets = 24 s. Afterwards, the analysis interval for the task phase was set to 1 s, starting from the moment each beep sound was played. Data from the first and last beep sounds in each set of 10 provided beeps were excluded from analysis. Consequently, the analysis time for each participant’s task phase was 1 s × 8 times × 3 sets, totaling 24 s (Figure 1); then, the RMS was calculated for each analysis interval using the following equation (1).

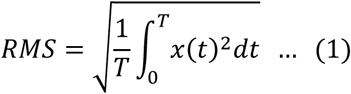

Here, *T* denotes the length of the analysis interval, and *x*(*t*) represents the EMG amplitude at time *t*. The RMS value for the resting phase was the minimum value from the 3 sets, while the RMS value for the task phase was the average of the three sets used for analysis. Furthermore, the RMS value for the resting phase was confirmed to be below 15 μV, the resting baseline [37]. A higher RMS value indicates stronger muscle activity.

Subsequently, using the RMS values for the resting and task phases calculated in the above analysis interval, the degree of pMA for the right extensor and right flexor was calculated using the following equation (2) [38].

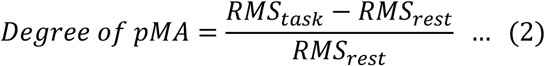

A degree of pMA of 0 indicates that the muscle activity of the right extensor or right flexor was the same during rest and during the ball-rolling task with the left hand. Furthermore, a higher degree of pMA indicates that the activity of the right extensor or right flexor was higher during the ball-rolling task with the left hand compared with the rest performance, indicating a stronger occurrence of pMA.

CMC was calculated during the task phase using the Electromagnetic Source Estimation Suite software (Cortech Solutions, Wilmington, NC, USA). For each EEG-EMG pair, coherence spectra were estimated using a fast Fourier transform-based spectral estimation method with 1-s windows and 0% overlap. CMC was calculated according to the following equation (3) for each EEG-EMG pair.

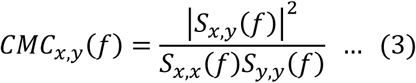

Here, *CMC_x_*_,*y*_(*f*) indicates the CMC value at frequency *f* for EEG or EMG electrodes *x* and *y*. Furthermore, *S_x_*_,*y*_(*f*) represents the cross-spectrum between electrodes *x* and *y* at frequency *f*, while *S_x_*_,*x*_(*f*) and *S_y_*_,*y*_(*f*) represent the respective auto-spectra of electrodes *x* and *y* at frequency *f*. *CMC_x_*_,*y*_(*f*) takes values between 0 and 1, with higher values indicating greater functional connectivity between *x* and *y*. We calculated the peak CMC value in the gamma frequency band (33–52 Hz) [16,39], which is involved in dynamic motor control, from CMC values computed at 1 Hz intervals. Because the frequency at which corticomuscular coupling is most pronounced may vary across individuals and EEG-EMG pairs, the maximum CMC value within this predefined gamma band was used as the peak gamma-band CMC. This approach enabled us to capture the strongest task-related corticomuscular coupling while avoiding post hoc selection of an arbitrary frequency range.

### Statistical analysis

First, we performed the Shapiro–Wilk test on all data to verify normality. In addition, we conducted a chi-square test to examine sex ratio imbalances between the young and older adults. A two-way ANCOVA was performed for minimum RMS in the resting phase, average RMS in the task phase, and the degree of pMA. Sex was used as a covariate, with muscle (extensor, flexor) and group (young adults, older adults) as the main factors. For all ANCOVA results, a Bonferroni post-hoc test was conducted for factors showing significant interactions or main effects.

Spearman’s partial correlation analysis, with sex as a covariate, was used to examine the relationship between the degree of pMA and CMC within each group. Specifically, Spearman’s partial correlation analysis was performed for the degree of pMA (right extensor) with respect to the CMC between EEG and extensor EMG, and for the degree of pMA (right flexor) with respect to the CMC between EEG and flexor EMG. Statistical analyses were performed using SPSS ver 29.0 (IBM Corp., Armonk, NY, USA) and Python 3.13.3 (Python Software Foundation, Wilmington, DE, USA). The statistical significance level was set at 5%.

## Data Availability

The datasets generated and/or analyzed during the current study are not publicly available because of privacy and ethical restrictions but are available from the corresponding author on reasonable request and with permission from the relevant ethics committee, where applicable.

## Acknowledgements

The authors express their sincere gratitude to all volunteers who participated in this study and to the staff who assisted with data collection.

## Author contributions

Conceptualization: SS and HN; Data curation: SS and HN; Formal analysis: SS and HN; Funding acquisition: SS and HN; Investigation: SS, SM, NS, SF, RY, TN, YS, and HN; Methodology: SS and HN; Project administration: SM and HN; Resources: SM and HN; Supervision: HN; Validation: SS, SM, NS, SF, RY, TN, YS, and HN; Visualization: SS and HN; Writing – original draft: SS and HN; Writing – review & editing: SS, SM, NS, SF, RY, TN, YS, and HN.

